# Improved diagnostic identification of urothelial carcinoma through solid-state nanopore determination of urinary hyaluronan size distribution

**DOI:** 10.64898/2026.08.12.26360208

**Authors:** Dorothea A. Erxleben, Suruchi Poddar, Carla M. Rodriguez, Paul H. Williams, Matthew A. Davis, Ronald L. Davis, Dixy E. Green, Paul L. DeAngelis, Elaheh Rahbar, Ekaterina S. Khvatkova, Carl D. Langefeld, Adam R. Hall

## Abstract

Urothelial carcinoma (UC) is among the most common malignancies worldwide and is known to exhibit a high recurrence rate. The relative lack of validated, non-invasive biomarkers for the disease challenges early detection and negatively impacts patient outcomes. The linear polysaccharide hyaluronan (HA) has been recognized as a potential source of diagnostic information for UC, with its urinary concentration shown to be predictive of disease severity. Here, we use solid-state nanopore (SSNP) sensing to investigate the value of urinary HA size distribution as an independent and complementary predictor of UC. We show that, when combined with urinary concentration, HA size distribution provides a significant improvement to the differentiation of healthy individuals from those with urinary tract diseases in general (AUC = 0.91, *p* < 0.05), as well as differentiation of individuals with UC from those without (AUC = 0.87, *p* < 0.05). These results establish the potential of SSNP-based HA profiling for non-invasive diagnostics of UC.

## Introduction

Urinary tract cancers include cancers of the urethra, bladder, ureter, and kidney (renal pelvis) and have both a high prevalence and a high mortality in the United States^1^ and globally^2^. The vast majority of these cancers are urothelial carcinomas (UCs), wherein the transitional epithelial cell lining within the urinary tract undergoes malignant transformation to result in either well-confined tumors (non-muscle invasive) or tumors that spread to the underlying muscle tissue (muscle invasive). The treatment of UC is guided by disease stage, with non-muscle invasive cancers typically managed through transurethral resection and muscle-invasive cancers often requiring more aggressive interventions that can include radical cystectomy and systemic chemotherapy or immunotherapy. While the 5-year survival rate for early-stage UC is generally high (for example, 98% for *in situ* bladder UCs and 73% for localized bladder UCs), late-stage survival can be extremely low, reducing to < 10% overall for metastatic forms^3^. This dichotomy underscores the importance of early identification for improving patient outcomes and, together with the frequent need for post-treatment recurrence monitoring, highlights the need for robust diagnostic strategies^4^. Unfortunately, available techniques for early screening of UC have significant limitations. For example, cystoscopy with biopsy is considered the gold-standard detection method^5^ but it is invasive, laborious, and requires analysis by a trained pathologist. Approaches like urinalysis and cytology hold potential for non-invasive monitoring but have exhibited high false positive rates and low sensitivities for early-stage lesions^5^. To fill this gap, urinary molecular biomarkers have been identified, including liquid biopsy panels (*e*.*g*., cell-free DNA^6^) and protein-based tests^7^. However, these assays exhibit limited sensitivity and specificity, especially in detecting early-stage UC^8^.

A molecular target that has generated significant consideration as a UC biomarker is the glycosaminoglycan hyaluronan (hyaluronic acid; HA), which is a key component of the extracellular and pericellular matrices and is known to play critical roles in cancer progression. For example, HA modulates immune evasion in the tumor microenvironment by forming a protective layer around cancer cells and engaging CD44 receptors to activate pro-tumor signaling pathways^9,10^. Tumor progression is also associated with upregulated expression of hyaluronidase enzymes that degrade HA, resulting in the accumulation of HA fragments in UC tissues^11,12^ that can subsequently induce angiogenesis^13–15^, stimulate cell proliferation^16^, and influence important activities like tumor cell adhesion and migration^11^. Because the urinary tract epithelium contacts the urine directly, these enhanced degradation dynamics can also result in a buildup of HA fragments in the urine^17^. Such an accumulation has been observed both in bladder cancer^14^ and in the context of residual UC tumors^18,19^ and has been correlated with metastatic potential^9,14^. But while urinary HA concentration has shown potential for non-invasive diagnostics, its power to classify patients by disease stage or to distinguish UC from benign disease is limited^14,18,19^. Consequently, there is a clear need for additional metrics that could yield improved predictive efficacy.

One important candidate for this purpose is the size distribution of HA, which has been reported to change in wound healing and disease states^20,21^ as a result of both altered HA metabolism (*e*.*g*., hyaluronan synthase and/or hyaluronidase enzyme activity) and increased fragmentation induced by reactive oxygen species associated with inflammatory processes^13^. Because the HA degradation processes that lead to urinary HA accumulation in UC may also be expected to impact its size, molecular weight (MW) distribution analysis could provide information that is complementary to HA quantitation alone. Established approaches for HA MW analysis have included mass spectrometry^22,23^, size exclusion chromatography coupled with multi-angle light scattering^24^, and gel electrophoresis^25^ but these methods have significant constraints that limit their utility for clinical investigations^26^, including complex instrumentation, restricted dynamic range, or high mass requirements – the last of these being especially challenging for an HA-sparse biofluid like human urine^27^.

Solid-state nanopore (SSNP) analysis addresses these constraints in a platform consisting of a single nanometer-scale aperture in a thin, insulating membrane through which HA molecules can be translocated electrically and assessed individually by their impact on the trans-membrane ionic current measured during threading^28,29^. We have previously demonstrated that the SSNP approach enables quantitative determination of HA MW distributions from as little as 10 ng total HA mass and with a size resolution at least as low as 50 kDa^30,31^.

Here, we investigate the potential predictive power of SSNP-determined urinary HA MW distribution as a UC biomarker, both alone and in combination with the existing metric of urinary HA concentration. We focus on urinary HA derived from four clinical cohorts: *(i)* healthy controls, *(ii)* individuals with other non-malignant diseases of the urinary tract, *(iii)* individuals with non-muscle invasive UC, and *(iv)* individuals with muscle invasive UC. After validating HA concentration differences between cohorts, we use calibrated SSNP analyses to ascertain an HA MW distribution for each urine specimen. We first compare all results through cumulative distribution analyses to identify group-level differences and then establish quantitative distributional metrics that can be used for binary sample classifications, improving differentiation significantly over demographics alone, and showing synergistic predictive effects with conventional HA concentration assessment. Our results establish SSNP MW analysis of urinary HA as a non-invasive diagnostic approach with potential for translation to the clinic.

## Results and Discussion

### Subjects

Urine specimens were collected from a total of 209 consented subjects under protocols approved by the Atrium Health Wake Forest Baptist Institutional Review Board (IRB nos. 00039804 and 00062432), including 52 healthy subjects (Cohort *H*, ages 20–82 years; median 74 years), 51 individuals with other non-malignant disease (Cohort *O*, 36–88 years; median 68 years), 56 individuals with non-muscle invasive UC (Cohort *N*, 48– 88 years; median 74 years), and 50 individuals with muscle invasive UC (Cohort *I*, ages 48–85 years; median 71 years). Subject age distributions are provided graphically in **Supplementary Figure S1**.

The demographic makeup of subjects in the H cohort was: 87% White, 10% Black or African American, and 4% Asian. Participants in this cohort were required to be over 18 years of age and weigh more than 50 kg.

Exclusion criteria included: pregnancy or active breastfeeding; fever, cough, or shortness of breath within the 7 days preceding collection; cold or flu within the 2 weeks preceding collection; taking of aspirin, NSAIDs (*e*.*g*., ibuprofen), acetaminophen, or any other anti-platelet or anti-coagulant drug within the 24 hours preceding collection; diagnosis of COVID within the 2 weeks preceding collection; providing more than two blood draws within the 7 days preceding collection; diagnosis of rheumatoid arthritis; diagnosis of cancer; diagnosis of fibromyalgia; diagnosis with cirrhosis of the liver; receipt of HA treatment (*e*.*g*., topical or injected HA) within the 2 months preceding collection; and active prescription of heparin intravesical therapy or Elmiron® (pentosan polysulfate sodium) for interstitial cystitis.

The demographic makeup of subjects in the urothelial disease cohorts (O, N, and I) was: 83% White, 11% Black or African American, 1% Asian, and 5% Unknown or Not Specified. Participants in these cohorts were required to be at least 18 years of age and within the 2 years prior to collection to have (1) received a clinical diagnosis or recurrence of UC or (2) been diagnosed with a non-malignant disease of the urinary tract (*e*.*g*., kidney stones). Exclusion criteria included: pregnancy; diagnosis with fibromyalgia; diagnosis with cirrhosis of the liver; diagnosis of rheumatoid arthritis; receipt of HA treatment (*e*.*g*., topical or injected HA) within the 2 months preceding collection; and active prescription of heparin intravesical therapy or Elmiron® (pentosan polysulfate sodium) for interstitial cystitis.

### Urinary HA Concentration

To corroborate the differences in urinary HA abundance reported in past literature^12,14,18,19,32^, we first assessed the concentration of HA in each urine specimen. For a comprehensive HA concentration assessment, we used a competitive quantitative enzyme-linked immunosorbent assay (ELISA) selected for its extended size-sensitivity for HA^33^ and while there is no perfect normalization method to account for subject hydration, we followed the precedent established for UC elsewhere^34,35^ and normalized HA concentration against urinary creatinine (UCr) content. Our measurements across all samples showed a general trend toward higher urinary HA concentrations with disease severity (**Figure 1**) and statistically-significant differences between groups (**Supplementary Table S1**) confirmed the utility of the metric as an indicator of urinary tract health, as in prior literature^14^. For example, HA concentration was elevated significantly (*p* < 0.05, adjusted for age and sex) for O, N, and I cohorts (*i*.*e*., all urothelial disease cohorts) relative to H controls. But particularly noteworthy were the highly-significant (*p* < 0.0001) differences observed between the H and O cohorts since these groups have been reported to be indistinguishable in past work^18^. Possible explanations for this distinction include dissimilarities in the specific balance of conditions represented among subjects in our O cohort as well as the narrower MW sensitivity^36^ of the non-competitive ELISA used in the previous quantification^18,19^. Significant differences were also identified between the N and I groups, in alignment with prior reports associating more advanced UC with increased HA concentrations^14^. In our analyses, the only groups that could not be differentiated readily were the O and N cohorts; this case may have been due to the high levels of inflammation associated with the specific pathologies represented among the non-malignant disease cohort, including kidney stones^16^, which may be more acute than some instances of non-muscle invasive UC.

**Figure 1.**
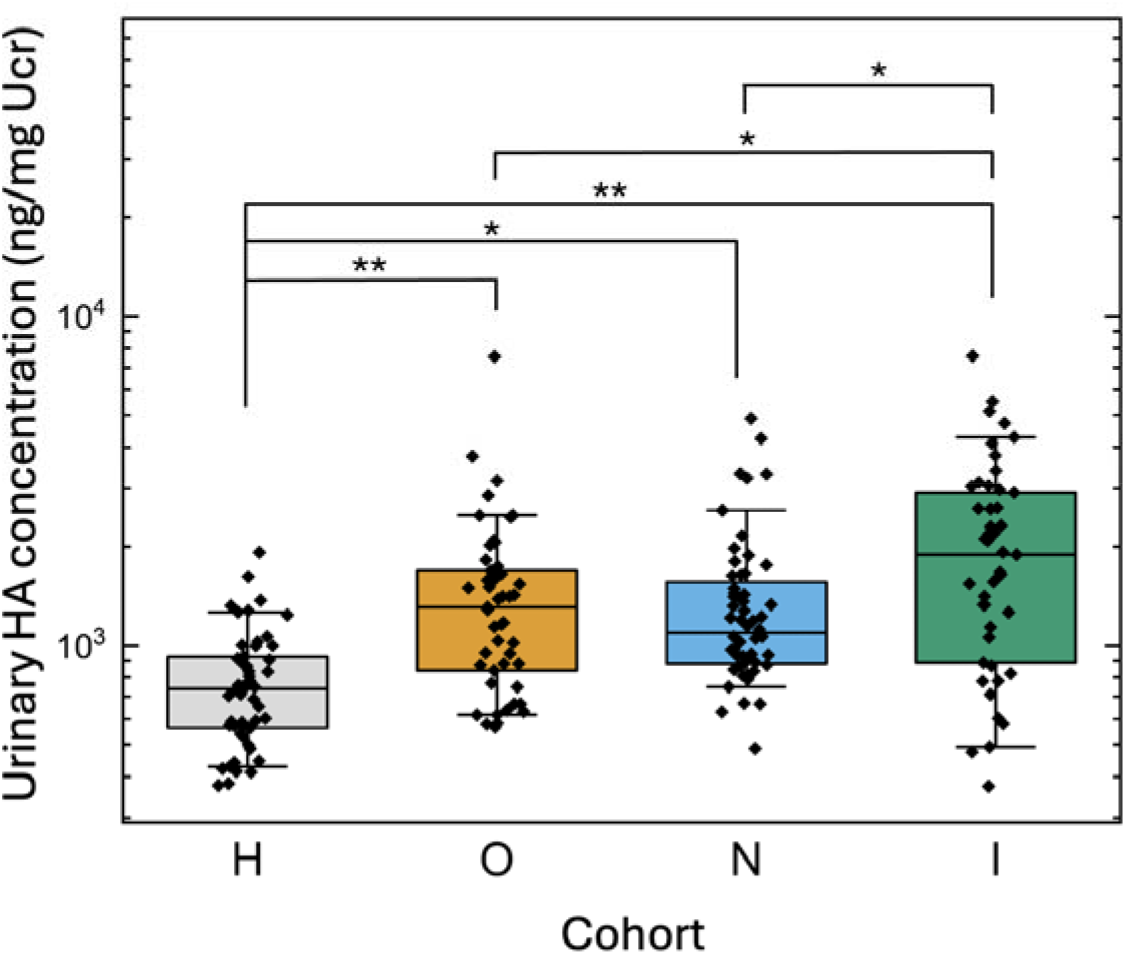
Urinary HA concentrations, normalized to UCr, measured across all four patient cohorts: healthy (H), other non-malignant diseases of the urinary tract (O), non-muscle invasive UC (N), and muscle invasive UC (I). Significance (adjusted for age and sex) are indicated by * (*p* < 0.05) and ** (*p* < 0.0001). Boxplots indicate the median (internal horizontal line), the 25^th^-75^th^ percentile range (box), and the 10^th^-90^th^ percentile range (whiskers). *N* = 51 (H), 49 (O), 56 (N), and 49 (I).

### Urinary HA MW Distribution

Having confirmed in our clinical specimens the prior findings regarding urinary HA concentration in the context of UC, we continued our investigation by probing HA MW distributions as an independent biomarker. Here, we first used an established extraction protocol^30,37^ with slight modifications (*Materials & Methods*) to isolate total HA from aliquots of the same urine specimens as above and then assessed them directly by SSNP (**Figure 2a**). For all extracted HA samples, the electrical signature of each translocated HA molecule – its event charge deficit (ECD, **Figure 2a**, inset) – was converted to a MW using an internal measurement calibration strategy designed to minimize device-to-device variation^26^ (*Materials & Methods*) and yielded for each urine specimen a unique distribution that captured its distinct HA MW profile (**Figure 2b**).

**Figure 2.**
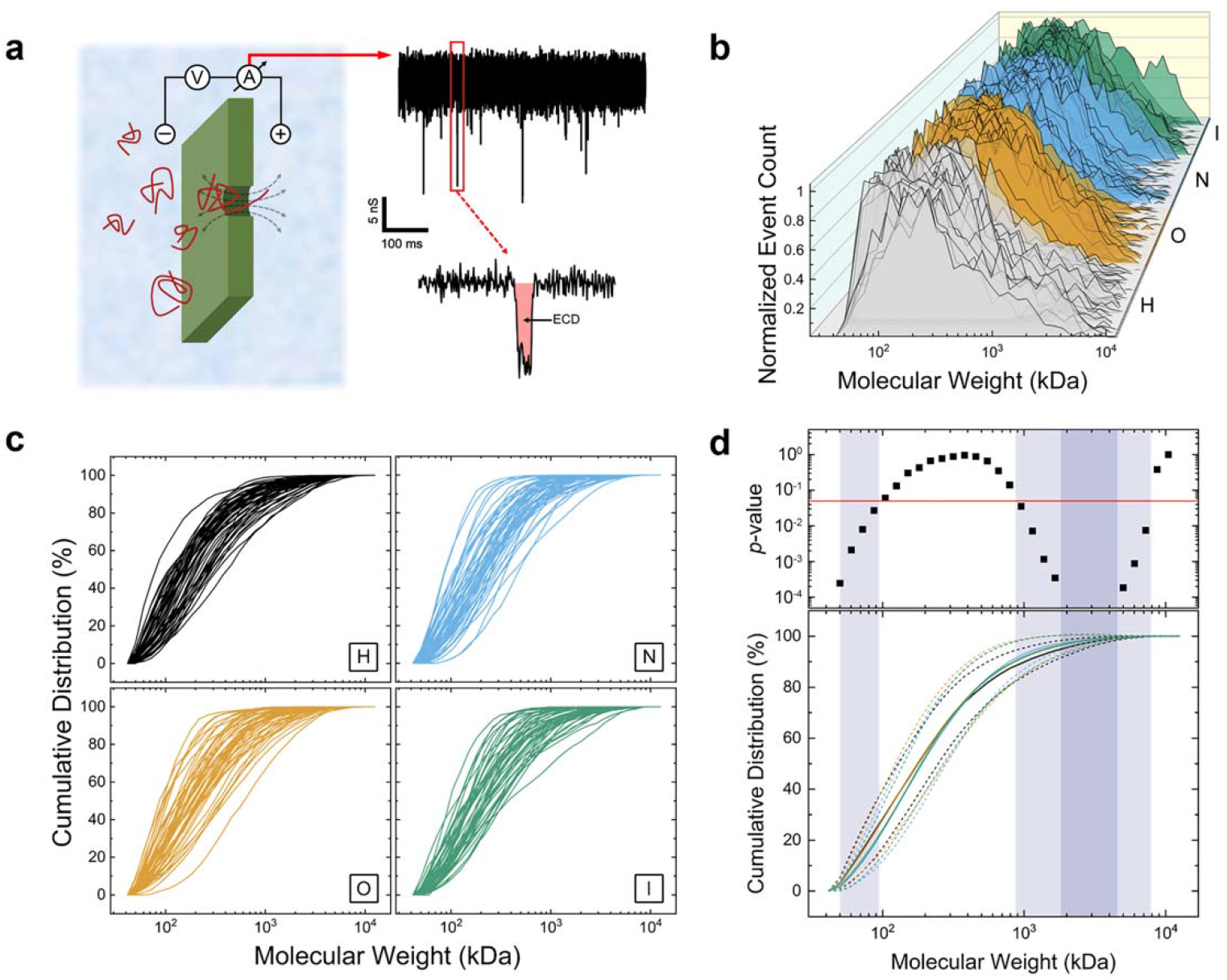
SSNP measurements of urinary HA and resulting distributional size analyses. **(a)** Schematic of the measurement approach wherein a voltage is applied across a single SSNP, causing electrical translocations of HA molecules (left). Resulting ionic current traces (right, top) feature temporary blockages (events), the area (or ECD, shaded region) of which can be correlated to MW. **(b)** Normalized HA size distributions across urinary tract disease states measured via SSNP. **(c)** Cumulative size distribution functions for all measured samples, divided by cohort. **(d)** Bottom: mean cumulative distribution functions for each cohort with dashed lines indicating their standard deviations (colors match (c)). Top: global *p*-values (by bin) resulting from a K-W test. Vertical shaded regions designate ranges of K-W significance, with light grey signifying *p* < 0.05 (indicated by the red horizontal line) and dark grey signifying *p* < 0.0001. *N* = 49 (H), 50 (O), 54 (N), and 48 (I).

Given the complexity of these physiological MW distributions, no clear qualitative differences were obvious between specimens or groups. We therefore proceeded to conduct statistical comparisons between the cohorts. We first constructed a cumulative frequency curve for each urine specimen (**Figure 2c**), indicating the fraction of molecules at or below each MW across the entire dynamic range of SSNP analysis (50 kDa–10 MDa) and providing a visual representation of how HA distributions evolved as a function of MW. To evaluate differences between groups, we employed the Kruskal-Wallis (K-W) test. This test is a nonparametric analogue of the one-way ANOVA for comparing more than two groups and was suitable for our distributions since they did not satisfy requirements of normality. The K-W test was used across the entire MW range to calculate the likelihood that cohort medians within each bin were statistically different. By considering the resulting *p*-values (**Figure 2d**, top) we observed significance (*p* < 0.05) in both the low-MW (50–95 kDa) and high-MW (869–7924 kDa) regions (**Figure 2d**, bottom, shaded regions), indicating that the underlying size distributions within the four cohorts were not equivalent within those MW bins.

While a K-W test could provide a global evaluation of differences across groups, it could not indicate which specific groups differed from each other. Consequently, the observation that significant distinctions existed in our data motivated the incorporation of quantitative MW distributional metrics into our analyses. For this, we employed the first four statistical moments of log_10_(MW) values of each distribution by calculating its *(i)* geometric mean (center), *(ii)* variance (spread), *(iii)* skewness (asymmetry), and *(iv)* kurtosis (tailedness) and then performed statistical comparisons between cohorts (**Supplementary Table S2**) using a series of logistic regression models. Among these comparisons, we observed no significant differences between the groups when considering either the geometric mean (**Figure 3a**) or the kurtosis (**Figure 3d**). For the mean, a possible explanation for this result could be that dysregulated processes of HA synthesis and degradation in diseases of the urinary tract^13^ effectively offset each other, resulting in no detectable net change in population centers. For kurtosis, all measured MW distributions were found to be platykurtic, featuring broad peaks and light tails relative to a normal distribution (*c*.*f*., **Figure 2b**). This profile reflects the heterogeneous nature of physiological HA and, together with our finite dynamic range, may account for the statistical similarity of kurtosis values across samples.

**Figure 3.**
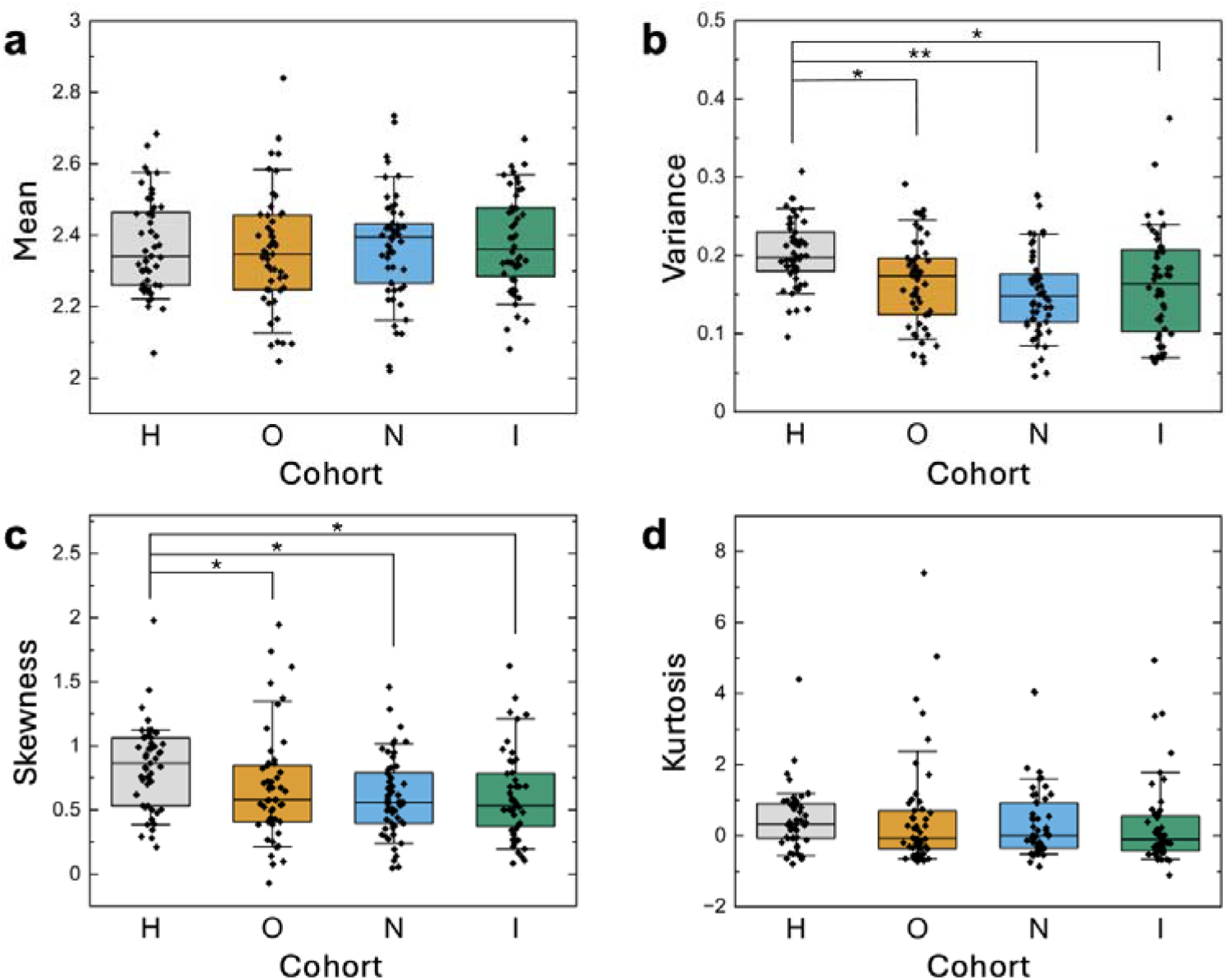
Moments of the log_10_(MW in kDa) distributions across patient cohorts, including **(a)** geometric mean, **(b)** variance, **(c)** skewness, and **(d)** kurtosis. Significant *p*-values (adjusted for age and sex) are indicated by * (*p* < 0.05) and ** (*p* < 0.0001). Boxplots present the median (internal horizontal line), the 25^th^-75^th^ percentile range (box), and the 10^th^-90^th^ percentile range (whiskers). *N* = 49 (H), 50 (O), 54 (N), and 48 (I).

In contrast, comparisons made using variance (**Figure 3b**) and skewness (**Figure 3c**) values revealed significant distinctions between cohorts (*p* < 0.05). Specifically, the healthy specimens differed significantly from all three disease cohorts, with the former exhibiting higher values than other groups for both metrics. We interpreted these differences as indicators of differential disease-related HA degradation profiles, with enhanced HA fragmentation in the O, N, and I cohorts leading to greater changes in the overall shapes of the measured distributions. The resulting significances suggested the value of incorporating variance and skewness as quantitative MW distribution metrics in subsequent analyses.

### Binary Classification Models

With the metrics available, we next performed a series of cumulative logistic regressions (**Supplementary Table S3**) to determine the classification efficacies of *(i)* age & sex, *(ii)* urinary HA concentration, and *(iii)* urinary HA MW distributional moments (variance & skewness). Only specimens with both urinary HA concentration and MW distribution data available were considered in these analyses. For robust comparisons, we calculated receiver operating characteristic (ROC) curves for the variables either alone or in combination. This enabled quantification of their relative performance as binary classifiers via calculation of the area under the curve (AUC) for each ROC, where a value of 1 is a perfect classifier with no false positives or false negatives. In conducting this analysis, we focused on two delineations of particular clinical diagnostic importance: healthy (H) versus all urothelial disease (O+N+I) and non-cancer (H+O) versus UC (N+I).

Since subject age and sex are known risk factors of UC^4,5^, we initially tested these as the only variables in our model. Our results corroborated that these purely demographic data alone had significant predictive power for both classifications, yielding an AUC value of 0.74 for the ROC curve comparing healthy with all disease cohorts (**Figure 4a**, black) and an AUC value of 0.78 for the ROC curve comparing non-cancer cohorts with cancer cohorts (**Figure 4b**, black). These baseline values could subsequently be used to evaluate the relative improvements provided by analytical urinary HA data. With the inclusion of UCr-normalized HA concentration (see **Figure 1**) along with patient demographics in the models, we found that the AUCs increased to 0.84 for healthy versus all disease (**Figure 4a**, purple) and to 0.81 for non-cancer versus cancer (**Figure 4b**, purple).

**Figure 4.**
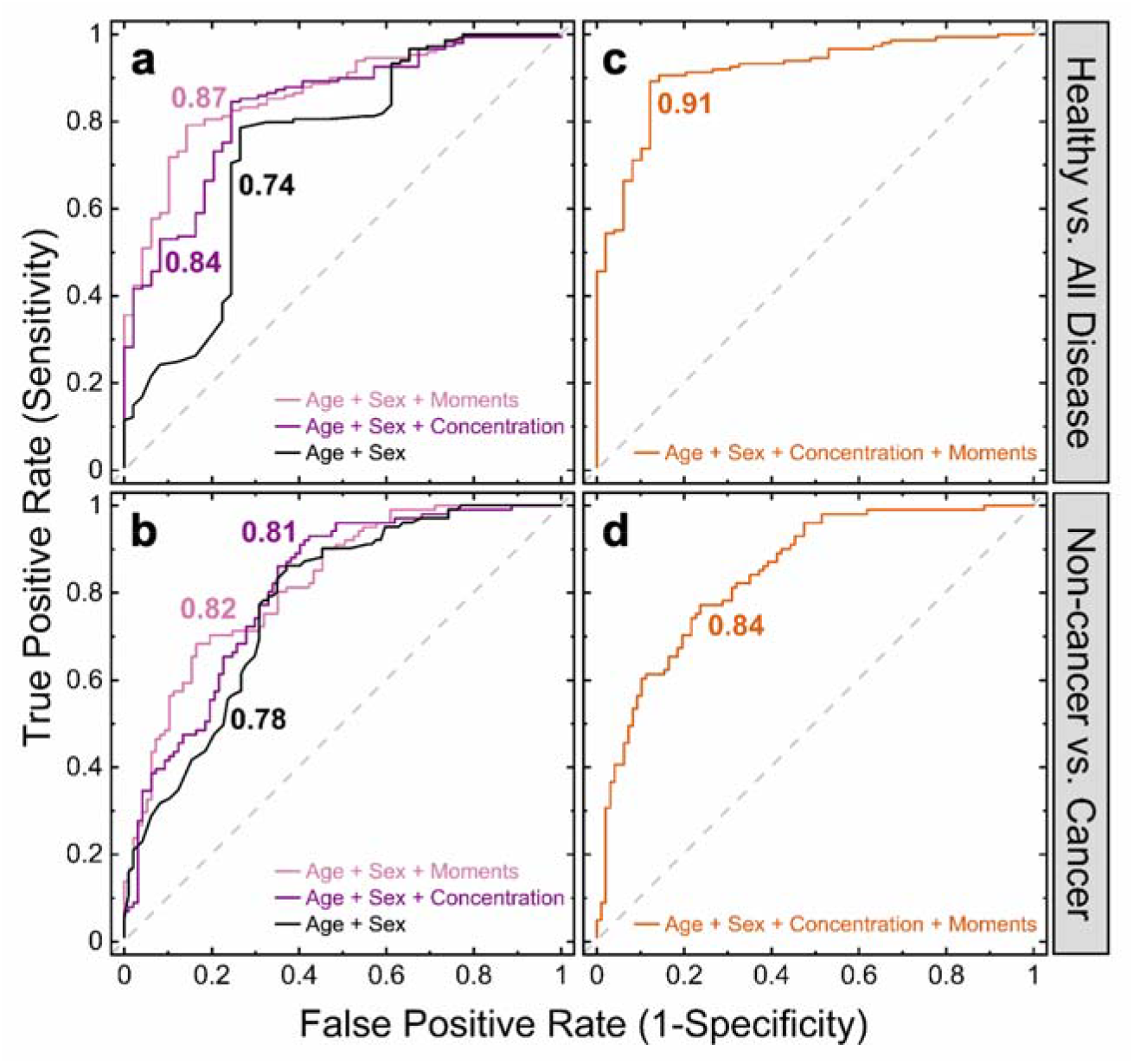
ROC curves for the logistic regression binary classification models differentiating cohort H from cohorts (O+N+I) (healthy versus all disease, **top**) and cohorts (H+O) from cohorts (N+I) (non-cancer versus cancer, **bottom**). The black line considers patient age & sex only. The purple line and the pink line add considerations of UCr-normalized HA concentration and the variance & skewness HA MW distributional moments, respectively. The dark orange line considers all four metrics collectively. AUCs are indicated on plots (color-matched). *N* = 49 (H), 48 (O), 54 (N), and 47 (I).

These improvements in discriminatory power were in line with prior literature^14,18,35,38^ and further confirmed the value of urinary HA abundance as a UC biomarker. By instead including the variance and skewness of the urinary HA MW distributions into the predictive models along with subject age and sex (**Figure 4a and b**, pink), we found AUC values of 0.87 and 0.82 for healthy versus all disease and non-cancer versus cancer, respectively. The increases in both values relative to the demographic-only baseline were larger than those associated with ELISA quantification, indicating that the two SSNP-derived HA MW distributional moments could provide classification that was superior to that of HA abundance. Finally, because the UCr-normalized urinary HA concentration and the MW distribution of urinary HA are likely to contain orthogonal information, we also evaluated a combined model that utilized subject age and sex along with both the HA abundance and the variance and skewness of the MW distributions. Here, the AUCs achieved higher values still, reaching 0.91 for healthy versus all urinary tract disease (**Figure 4c**) and 0.84 for non-cancer versus cancer (**Figure 4d**).

These findings indicated that joint consideration of HA concentration along with the MW distribution variance and skewness could classify urine samples by disease state better than either metric alone.

While our findings render a potentially powerful tool for assessing urinary tract pathology in general, the predictive power for UC specifically was somewhat less robust, yielding only a modest improvement when combining all metrics into a single model. Therefore, we finally sought out an additional metric that could improve the efficacy of the non-cancer versus cancer classification. For this, we returned to the results of the K-W analysis that indicated global differences among the four cohorts within specific size ranges of the urinary HA cumulative frequency curves across the dynamic range (see **Figure 3d**, shaded regions). To investigate whether bin-level data could improve discriminatory accuracy further, we focused on the two ranges of MW in which significant global differences (*p* < 0.05) were observed: one low-MW (50–95 kDa) and one high-MW (869–7924 kDa) range. We employed the Fisher’s Protected Least Significant Differences (LSD) test to evaluate bin-level pairwise differences, and through these analyses we identified several bins for which the measured values differed significantly between cohorts (**Figure 5a**). For example, numerous bins showed distinctions between H and either the N or I cohort and additional significant differences were found to occur exclusively in the high-MW range (between O and H) and exclusively in the low-MW range (between O and I).

**Figure 5.**
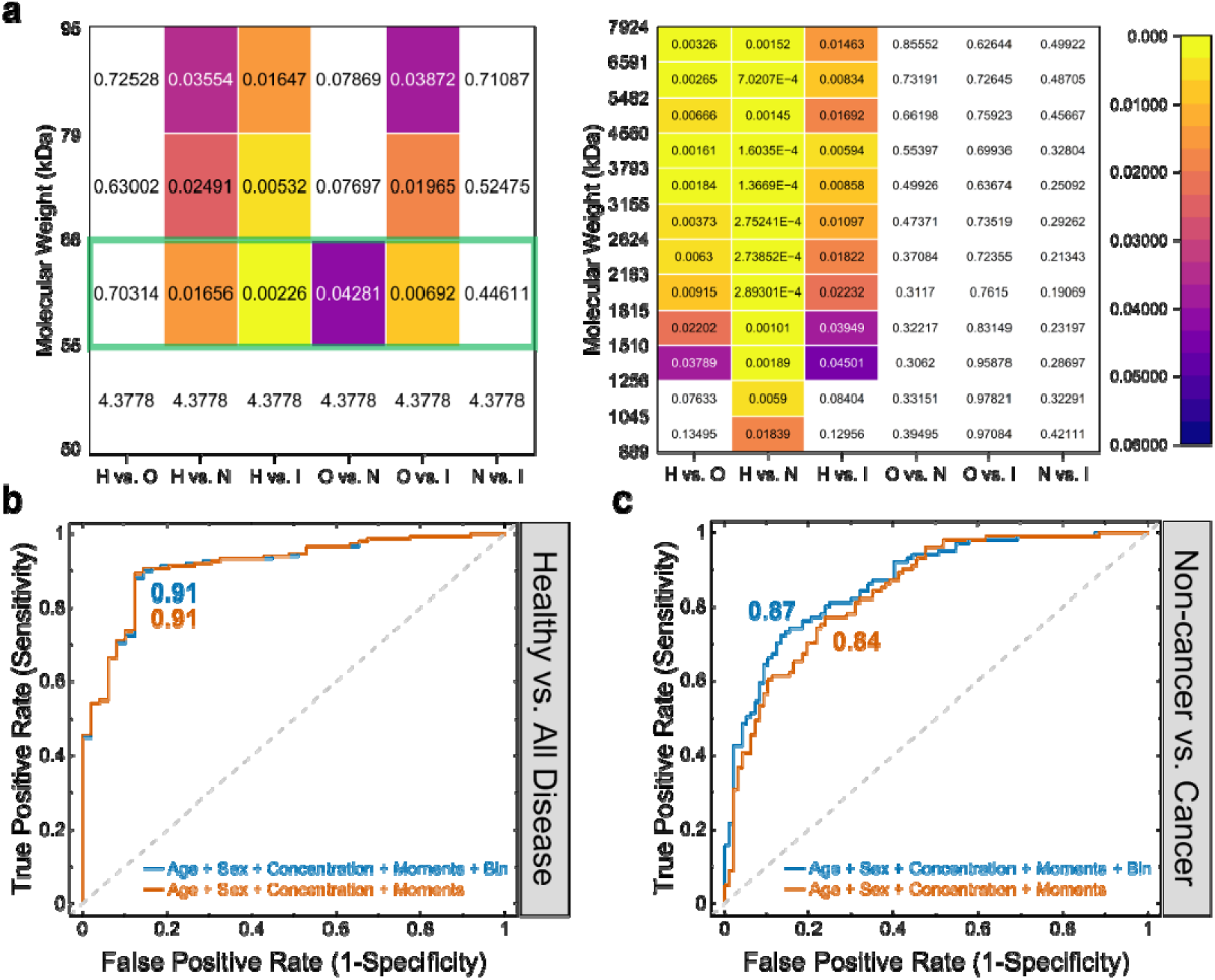
Consideration of MW bin-level differences among cohorts. **(a)** Heatmaps showing bin-level *p*-values resulting from Fisher’s LSD pairwise comparisons of the low-MW (left) and high-MW (right) ranges. ROC curves for the logistic regression binary classification models differentiating **(b)** cohort H from cohorts O, N, and I (healthy versus all disease) and **(c)** cohorts H and O from cohorts N and I (non-cancer versus cancer). The dark orange lines consider patient age & sex, UCr-normalized urinary HA concentration, and variance & skewness HA MW distributional moments (data also shown in **Figure 4c** and **d**). The dark blue line considers all of these metrics plus the cumulative frequency at the 55–66 kDa bin. AUCs are indicated on plots (color-matched). *N* = 49 (H), 48 (O), 54 (N), and 47 (I).

Unique among all analyses in this report, we identified a low-MW bin (55–66 kDa) that yielded statistical significance between the O and N cohorts (**Figure 5a**, green box; **Supplementary Figure S2**). Because the distinction between O and N was critical for the differentiation of non-cancer from cancer, we finally incorporated this bin-level quantitative descriptor of the cumulative HA MW distribution function into our model to test its impact on efficacy. For this process, we reconstructed ROC curves for each of our two binary classifications by adding the 55–66 kDa bin cumulative frequency curve values as an independent metric in addition to subject demographics (age and sex), UCr-normalized HA concentration, and the variance and skewness of the urinary HA MW distributions. In comparing healthy versus all others (**Figure 5b**), we found that this combined model yielded a ROC curve with an AUC of 0.91. This value was identical to the results of the model without the inclusion of bin-level data and indicated that its consideration was unnecessary for the classification. However, for the non-cancer versus cancer model (**Figure 5c**), collective consideration of all metrics yielded a ROC curve with an increased AUC of 0.87. This improvement over the prior results that excluded the cumulative MW distribution bin data (AUC=0.84) demonstrated the value of the alternative distributional metric to the model and brought the non-cancer versus cancer classification closer to unity with the healthy versus all disease classification above.

## Conclusion

Urinary HA concentration is known to increase in individuals with UC, scaling with severity to an extent, but the effectiveness of this metric as an independent biomarker for the disease has been limited. While the same degradation mechanisms that can cause HA accumulation in the urine may also be expected to impact its size distribution, conventional technologies for measuring HA MW have lacked sufficient sensitivity to enable its regular assessment, especially in a typically HA-sparse biofluid like urine. In this study, we have addressed this challenge through the application of SSNP sensing: a promising analytical tool capable of HA size determination across a broad MW range and with unparalleled sensitivity. Using the technique to probe urinary HA from human subjects across four cohorts (healthy, other non-malignant disease, non-muscle invasive UC, and muscle invasive UC), we demonstrated that two statistical moments of each MW distribution (variance and skewness) could be used to make binary distinctions between clinically-relevant classification groups, including differentiating healthy from all disease and non-cancer from cancer. Our results indicated that the performance of the selected distributional moments was better than that of HA concentration for the two binary classifications and, critically, that consideration of both metrics together improved the predictive power of the model compared to patient demographics more than either metric independently, establishing their orthogonality as biomarkers. Finally, we incorporated bin-level data into the model by considering an HA MW cumulative frequency curve bin (55–66 kDa) with key significance between cohorts O and N. In total, we achieved a high accuracy for both healthy versus all disease (AUC = 0.91) and non-cancer versus cancer (AUC = 0.87) classifications.

Combined with the non-invasive nature of urinalysis in general, the quantitative efficacy of SSNP sensing for classifying patient specimens positions the approach as a potentially useful diagnostic tool for UC detection and staging. For example, early potential clinical applications could include screening of high-risk individuals, or recurrence monitoring for UC patients following surgical resection. To be useful in these and other clinical applications, it will be critical to test the replicability and generalizability of our findings in larger, multi-ethnic cohorts, and to determine how classification may be impacted by additional demographic profiles (*e*.*g*., race, body mass index, smoking status)^4,5^. While the median ages for all cohorts were comparable and our analyses adjusted for age where appropriate, the healthy cohort featured younger subjects and a wider variation in age due to the higher prevalence of UC and other non-malignant urinary tract diseases in older individuals. Follow on studies may seek to better age-match the specimens. Similarly, the particular types and severity of diseases that compose the other non-malignant cohort may be critical to differentiation and could be standardized more fully. Future work may also investigate HA populations in earlier and more distinct stages of UC progression to improve early-stage detection, especially since biomarkers for this disease type currently remain limited in their sensitivity and specificity^8^.

From a platform perspective, continuing advancements in the HA size resolution of SSNPs^31^ will also improve discrimination with the technique, with opportunity for improving the distinction between O and N groups in the lower MW ranges (< 50 kDa). In addition, ongoing developments in methods for HA extraction^37^ could help to streamline the overall process and address the need for standardized procedures^8^ in clinical translation.

Finally, because SSNP event rate can in principle be used to determine analyte concentration^29^, it may be possible to assess both the abundance and the MW distribution of urinary HA with the same platform to maximize classification accuracy in a single, unified measurement. Overall, our findings have established urinary HA MW analysis by SSNP sensing as a potentially powerful method for patient classification in the context of UC. By combining this approach with conventional concentration assessment, we achieved high diagnostic accuracy and demonstrated the value of the approach in conducting non-invasive disease detection and monitoring.

## Materials and methods

### Urine samples

Urine specimens were obtained in the clinic setting and immediately stored at 2–8 °C until transfer to the laboratory. Once collected, each specimen was gently inverted 10 times and then transferred into two 15 mL tubes and centrifuged at 1,000 ×*g* for 10 min at RT to remove particulates. The supernatants were recovered and pooled into a fresh 50 mL tube for storage at −80°C until further processing.

### Urinary HA concentration determination

HA concentrations were measured in triplicate using a commercial competitive HA ELISA (K-1200, Echelon) following manufacturer’s directions. Due to the heterogeneity of clinical urine samples, results with a coefficient of variation < 20% were deemed acceptable for use in subsequent analyses. Results below the limit of detection were reported as “0” in subsequent analyses. To account for patient hydration, HA concentrations were scaled against UCr concentration, measured in duplicate with a commercial colorimetric assay (DICT-500, QuantiChrom™) following the manufacturer’s directions. Prior to analysis, each specimen was vortexed to redissolve any storage-induced sedimentation and, if still unresolved, centrifuged at 2,000 ×*g* for one minute prior to supernatant collection for testing. Results with a coefficient of variation < 10% were considered acceptable for subsequent analyses.

### HA extraction

In preparation for SSNP MW analysis, HA was extracted from each urine specimen following an immunoprecipitation-like approach described elsewhere^29,30,37^ with some modifications. HA ELISA data (see above) was employed to determine sample volumes for extraction, targeting 9 µg of total HA when possible. This large mass was selected to ensure robust nanopore analyses across the dynamic range for all samples because ELISA quantification does not indicate the percentage of total HA with MW below the SSNP limit of detection (50 kDa). For specimens where this target mass could not be obtained from the available volume (as little as 660 ng total HA mass in the study), observed SSNP signals were still sufficient for MW analysis.

Each urine specimen was subjected to ultrafiltration (Amicon® Ultra Centrifugal Filters, 30K MWCO, Millipore Sigma), serving both to concentrate the varying sample volumes down to 150–300 µL and to exchange the buffer for 1X PBS in support of subsequent extraction steps. Each sample then underwent a broad-spectrum protease digestion (Proteinase K, Invitrogen) followed by treatment with phenol:chloroform:isoamyl alcohol (25:24:1) and centrifugation (14,000 ×*g*, 15 minutes, 20°C) in a phase-lock gel tube made in-house by adding approximately 400 mg of high-vacuum grease (Dow Corning) to a 2 mL microcentrifuge tube. The aqueous fraction (containing HA and other hydrophilic components) was recovered and stored at 2–8°C until further processing.

For selective HA extraction, superparamagnetic beads (Dynabeads™ M-280 Streptavidin, 11206D, Thermo Fisher Scientific) were conjugated with biotinylated VG1 (bVG1, G-HA02, Echelon Biosciences) as described elsewhere^30,37^. Phenol-extracted aqueous solution (150–300 µL, see above) containing HA was incubated with 1.5 mg of VG1 beads for at least 15 minutes^37^ at RT on a rotary mixer, after which the beads were collected magnetically to remove the capture solution and washed three times with 1X PBS. Bound HA was eluted by resuspending the beads in 40 µL of SSNP measurement buffer (6 M LiCl, 10mM Tris, 1 mM EDTA (pH 8)) for at least 15 minutes^37^ at RT on a rotary mixer and decanted under magnetic field to retain the beads. Resulting HA isolates in measurement buffer and recovered beads (saved for reuse^37^) were stored separately at 2–8°C until further use.

### SSNP analysis of HA MW

SSNP MW analyses were conducted on extracted HA as described in detail previously^30,31^ with some modifications. Devices consisting of a single pore in a low-stress silicon nitride thin-film membrane were either (1) fabricated by Helium ion milling^39^ (Orion Plus; Zeiss, Inc.) or controlled dielectric breakdown^40^ (Northern Nanopore Instruments, Inc.) in custom 20 nm thick membranes in a 10 µm x 10 µm window (Norcada, Inc.), or (2) fabricated commercially by wafer-scale silicon processing in 30 nm thick membranes in a 20 µm x 20 µm window (Norcada, Inc.). All SSNPs used for measurements displayed a linear current-voltage curve with resistance values indicating diameters in the range of 5–14 nm as calculated with an established model that assumes an effective pore thickness of 1/3 the total membrane thickness^41^. Prior to use, an SSNP was mounted into a custom 3D-printed flow cell (Carbon, Inc.) and measurement buffer (6 M LiCl, 10mM Tris, 1mM EDTA, pH 8.0) was introduced to the two chambers surrounding the membrane. Ag/AgCl electrodes were used to connect to a patch-clamp amplifier (Axopatch 200B, Molecular Devices) for electrical measurement.

HA eluate in measurement buffer (10 µL) was loaded on one side of the SSNP membrane and a 300 mV bias was applied as the trans-membrane current was monitored at a rate of 200 kHz using a 100 kHz four-pole Bessel filter. Data were collected and analyzed using a custom LabVIEW program (National Instruments) with which an additional 5 kHz low-pass filter was applied during analysis. Molecular translocations (events) were identified as temporary reductions in the measured ionic current and were considered for analysis only if (1) the maximum amplitude differed from the baseline by at least 5 standard deviations of the root-mean-square (RMS) noise and (2) the total event duration was at least 25 μs. For each translocation recorded, amplitude and duration were used to calculate an event area (event charge deficit^29^, or ECD) through which a MW could be assigned via comparison to a calibration curve. To maximize measurement precision, we implemented an internal standard approach in which a custom HA ladder (consisting typically of 111, 485, and 1076 kDa quasi-monodisperse HA standards^42^ or similar) was measured on each SSNP prior to sample analysis. This strategy enabled the construction of an ECD vs. MW calibration curve individualized for each device, mitigating variability in pore resolution. Only events corresponding to MWs between 50 kDa and 10 MDa were considered in the analyses. All log_10_(MW) distribution histograms were generated with a bin width of 0.08.

### Statistical analyses

For statistical analyses that used UCr-normalized urinary HA concentration as the outcome, data were transformed using a natural logarithm (*i*.*e*., log_e_(([HA]/[UCr])+1), which was selected to best approximate distributional assumptions of conditional normality and homogeneity of variance. This transformation did not alter inferences as compared to investigating data on a log_10_ scale, which is how we present our data graphically to maintain consistency across results and help guide visual interpretations. Patients with HA ELISA values below the assay limit of detection (50 ng/mL) were assigned a value of 0 before transformation.

Cumulative frequency curves were generated for each specimen (*c*.*f*., **Figure 2c**) using log_10_-transformed MW histograms with a bin width of 0.08. To evaluate the significance of differences between the cohorts, a K-W test (the non-parametric analogue to one-way ANOVA for comparing more than two groups) was run between the four cohorts across all bins. The first four distributional moments (mean, variance, skewness, and kurtosis) were also calculated for the log_10_(MW) distribution resulting from each specimen. To test for differences between disease groups, a linear regression model was computed with the four moments as the outcomes, adjusting for age, and sex as covariates. Pairwise differences were computed using the Fisher’s Protected LSD multiple comparisons approach for an individual log_10_(MW) bin as the outcome. Given the ordinal nature of severity of the disease groups, cumulative logistic regression models computed with variance and skewness as predictors and adjusting for age, sex, and UCr-normalized urinary HA concentrations enabled a quantitative assessment of the individual effects of each variable independently, adjusting for the above covariates and the other moment in the model (*i*.*e*., type III hypotheses). In this series of models where UCr-normalized HA concentrations were used as a predictor, data were transformed by a square root function to best match distributional assumptions.

## Data Availability

All data produced are available online at the Harvard Dataverse

https://doi.org/10.7910/DVN/EUMTTO

## Author contributions

D.A.E. performed method development and data collection, analyzed the data, contributed to experimental design, collected and processed samples, and wrote the manuscript. S.P. contributed to bead extractions and nanopore measurements. C.M.R. contributed to nanopore measurements. P.H.W. contributed to urinary creatinine measurements. M.A.D. contributed to ELISA measurements. R.L.D. oversaw clinical recruitment and provided biospecimens. D.E.G. and P.L.D. synthesized quasi-monodisperse HA standards. E.R. contributed to experimental design and aided in the collection of healthy urine specimens. E.S.K. and C.D.L. performed statistical analyses and contributed to manuscript preparation. A.R.H. oversaw the project, contributed to experimental design, and wrote the manuscript. All authors contributed to the editing and review of the manuscript.

## Acknowledgements

This work was supported by the National Institutes of Health (NIH) Grants R01 GM134226 (to A.R.H. and E.R.), P41 EB020594 (to A.R.H.), and K25 HL133611 (to E.R.). The authors thank Felipe Rivas Duarte for aiding in urine sample collection.

