## Supplemental Figures and Tables for "Improved diagnostic identification of urothelial carcinoma through solid-state nanopore determination of urinary hyaluronan size distribution"

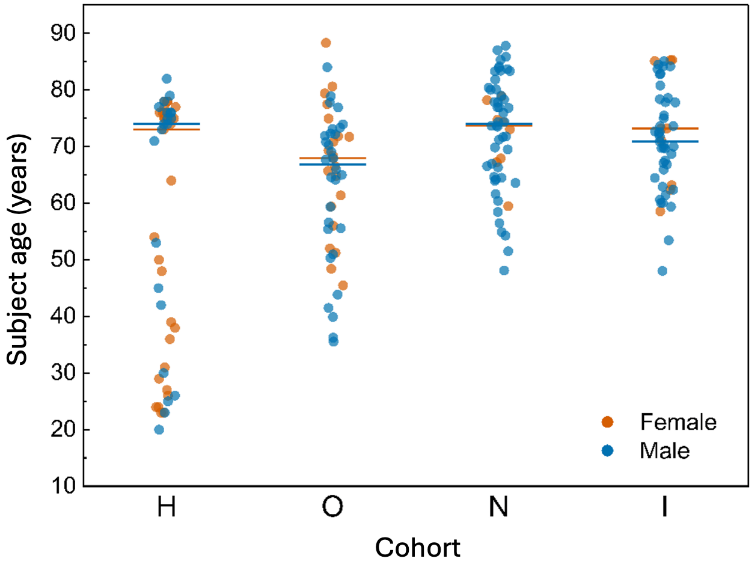


**Supplementary Figure S1.** Plotted subject demographics showing age and biological sex (orange represents females and blue represents males) of all subjects across the four cohorts including healthy (H), other non-malignant disease (O), non-muscle invasive UC (N), and muscle invasive UC (I). Horizontal lines represent median values (separated by biological sex, as indicated by color). **H**: *N* = 31 females (ages 23–78 years; median 73 years) and 21 males (20–82 years; median 74 years); **O**: *N* = 21 females (46–88 years; median 68 years) and 30 males (36–84 years; median 67 years); **N**: *N* = 8 females (ages 59–79 years; median 74 years) and 48 males (ages 48–88 years; median 74 years); **I**: *N* = 9 females (ages 59–85 years; median 73 years) and 41 males (ages 48–85 years; median 71 years).

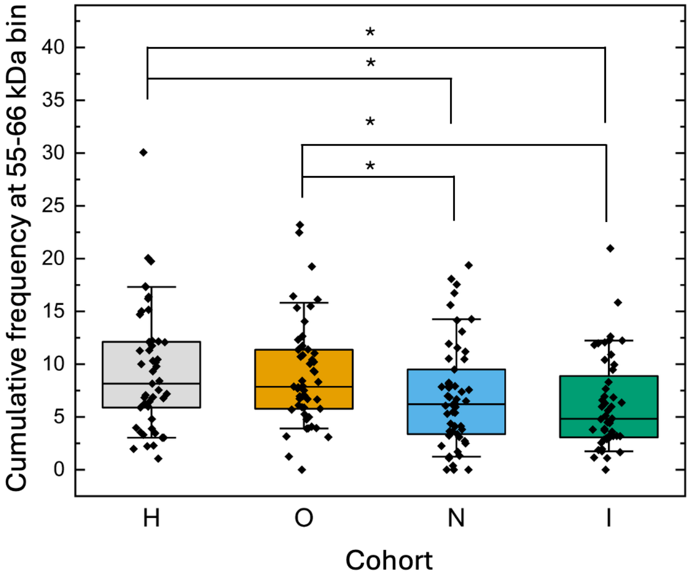


**Supplementary Figure S2.** Comparison of cumulative frequency values at the 55–66 kDa bin across patient cohorts including healthy (H), other non-malignant disease (O), non-muscle invasive UC (N), and muscle invasive UC (I). Significance ($p < 0.05$, adjusted for age and sex) is indicated by *. Boxplots present the median (internal horizontal line), the 25$^{th}$-75$^{th}$ percentile range (box), and the 10$^{th}$-90$^{th}$ percentile range (whiskers). $N$ = 49 (H), 48 (O), 54 (N), and 47 (I).

| **Cohort comparison** | ***p*-value** |
|---|---|
| H vs. O | < 0.0001 |
| H vs. N | 0.0005 |
| H vs. I | < 0.0001 |
| O vs. N | 0.5819 |
| O vs. I | 0.0366 |
| N vs. I | 0.0055 |

**Supplementary Table S1.** Pairwise comparisons of creatinine-normalized urinary hyaluronan concentrations between cohorts.

| **Cohort comparison** | **Variance *p*-value** | **Skewness *p*-value** |
|---|---|---|
| H vs. O | 0.0020 | 0.0240 |
| H vs. N | < 0.0001 | 0.0020 |
| H vs. I | 0.0019 | 0.0038 |
| O vs. N | 0.1951 | 0.2483 |
| O vs. I | 0.7422 | 0.3478 |
| N vs. I | 0.3158 | 0.8332 |

**Supplementary Table S2.** Pairwise comparisons of the variance and skewness of molecular weight distributions between cohorts.

| Parameter | Standard Estimate | Wald Error | $\chi^2$ | *p*-value |
|---|---|---|---|---|
| Age | 0.0371 | 0.0103 | 12.8877 | 0.0003 |
| Sex | -1.5075 | 0.3132 | 23.1685 | < 0.0001 |
| $\log_{10}$(MW) Variance | -7.6768 | 2.4083 | 10.1613 | 0.0014 |
| $\log_{10}$(MW) Skewness | -1.5362 | 0.3912 | 15.4228 | < 0.0001 |
| sqrt([HA]/[UCr]) | 0.0608 | 0.0119 | 26.291 | < 0.0001 |

**Supplementary Table S3.** Multivariable logistic regression to test the independent association of variables with the cohorts. Analysis of Maximum Likelihood Estimates for cumulative logistic regression of the study results, where *p*-values indicate the significance of each of the statistics independent of the others.
